# Retinal pigment epithelial cells reduce vascular leak and proliferation in retinal neovessels

**DOI:** 10.1101/2024.07.15.24306102

**Authors:** Simone Tzaridis, Edith Aguilar, Michael I Dorrell, Martin Friedlander, Kevin T Eade

## Abstract

Retinal pigment epithelial (RPE)-cells possess numerous functions and may respond to stress and damage of the neuroretina. In different neurodegenerative diseases, including age-related macular degeneration (AMD), retinitis pigmentosa, and macular telangiectasia type 2 (MacTel), RPE-cells have been shown to proliferate and migrate into the neuroretina, forming intraretinal pigment plaques. Though pigmentary changes are associated with disease progression, it is not known if their presence is protective or detrimental. In this study, we evaluated the impact of pigment plaques on vascular changes and disease progression in patients with macular telangiectasia type 2 (MacTel), an example of a progressive neurodegenerative retinal disease. We then studied underlying pathomechanisms using a mouse model mirroring these changes, the very-low-density lipoprotein receptor mutant (Vldlr–/–) mouse.

In a retrospective, longitudinal study, we analyzed multimodal retinal images of patients with MacTel and showed that pigment plaques were associated with a decrease in vascular leakage and stabilized neovascular growth. Using genetic approaches, we analyzed changes in expression levels of relevant genes in the RPE and retinas of Vldlr-/- mice during RPE-proliferation and migration. Our data indicated that RPE-cells transitioned from an epithelial to a mesenchymal state (“epithelial-mesenchymal transition”, EMT), proliferated and accumulated along neovessels. Using dextran angiography and immunofluorescence, we demonstrated that the perivascular accumulation of RPE-cells reduced vascular leakage. Pharmacologic inhibition of EMT led to a decrease in pigment coverage and exacerbation of neovascular growth and exudation.

Our findings indicate that the proliferation, migration and perivascular accumulation of RPE-cells may stabilize vascular proliferation and exudation, thereby exerting a protective effect on the diseased retina. We conclude that interfering with this “natural repair mechanism” may have detrimental effects on the course of the disease and should thus be avoided.

## Introduction

Retinal pigment epithelial (RPE)-cells possess numerous functions and may respond to stress and damage of the neuroretina.[1] In different neurodegenerative diseases, including age-related macular degeneration (AMD), retinitis pigmentosa, and macular telangiectasia type 2 (MacTel), RPE-cells have been proposed to proliferate and migrate into the neuroretina, forming intraretinal pigment plaques.[2-5] Increasing evidence suggests that during this process, RPE-cells transition from an epithelial to a mesenchymal state (“epithelial-mesenchymal transition”, EMT), granting these cells mesenchymal properties such as the ability to proliferate and migrate.[6, 7] Though pigmentary changes are commonly associated with disease progression, their role is not fully understood. This association could represent a causative relationship whereby the pigmentary changes contribute to disease progression, pigmentary changes could be protective from worsening disease, or pigmentation could simply be an effect of other disease-causing retinal changes that are unrelated to disease progression.

MacTel is an example of a progressive, neurodegenerative retinal disease that affects the central retina. Secondary vascular alterations are commonly observed.[8] Pigment plaques are a frequent finding in MacTel and pigmented changes and vascular alterations have been found to collocate.[3, 5] Early vascular changes of the disease include telangiectasia and increased vascular leakage, indicating a dysfunctional inner blood-retina barrier.[8] With disease progression, a shift of vessels from the deep retinal plexus to the outer retina as well as the formation of retinal-choroidal anastomoses and outer retinal neovascularization has been described. These changes may precede the formation of vision-threatening exudative subretinal neovascularization in some cases,[9-12] and the origin of neovessels has been ascribed to the retinal rather than the choroidal vasculature.[13-16] Recent imaging studies in MacTel proposed that the formation of outer retinal neovascularization induced proliferation of the RPE, upon contact between the RPE and outer retinal vessels.[17] It has been supposed that RPE cells then use these abnormal vessels as a scaffolding to migrate into inner retinal layers, where they form dense pigmented plaques.[5] Their role during disease progression and their impact on vascular changes, including vascular leakage and proliferation, have not yet been evaluated. We hypothesized that the perivascular accumulation of RPE-cells may stabilize vascular proliferation and reduce vascular permeability.

The very-low-density lipoprotein receptor (Vldlr) mutant (Vldlr–/–) mouse represents a common model of subretinal neovascularization, and is used to study disease characteristics of MacTel, retinal angiomatous proliferation and other conditions.[18, 19] As in MacTel, in Vldlr–/– mice, neovascular changes originate from the retinal vasculature. Retinal vessels proliferate, grow to the outer retina, and subsequently form subretinal neovascularization. As the disease progresses, mice show a proliferation of RPE cells with accumulation along neovascular tufts, followed by the migration of pigmented cells along retinal vessels into the neuroretina.[18, 20, 21] These events mirror the key neovascular and pigmentation-related events observed in MacTel and other diseases with subretinal neovascularization, making the Vldlr-/- mouse an ideal model to study these changes.

In this study, we aimed to (I) evaluate the role of pigment plaques on vascular changes and disease progression in MacTel, and (II) study underlying disease mechanisms using the Vldlr-/- mouse model mirroring these changes.

First, we show in a longitudinal, retrospective study of eyes with MacTel that perivascular pigment accumulation was associated with reduced vascular leakage and decreased *de novo* formation of exudative neovascularization. We then explored whether the observed associations were causally related and which mechanisms led to perivascular pigment accumulation in Vldlr-/- retinas. We found an enrichment of EMT-inducers and key mesenchymal markers in the RPE of Vldlr-/- mice. Pharmacologic inhibition of EMT-inducers led to decreased perivascular pigment accumulation and enhanced neovascular growth and exudation in the Vldlr-/- model, indicating a protective effect of pigmentary changes on vascular proliferation, mitigating vascular leak and proliferation. Based on our findings, we propose that EMT of the RPE, followed by proliferation, migration and perivascular accumulation may function as a “natural repair mechanism”, exerting beneficial, anti-angiogenic and anti-exudative effects on the diseased retina. As such, we conclude that interfering with these mechanisms may have detrimental effects on the course of the disease and should thus be avoided.

## Results

### Perivascular accumulation of pigment is associated with decreased vascular leakage in eyes with MacTel

To investigate the impact of perivascular pigment accumulation on vessel leakage and proliferation in MacTel, we compared the longitudinal courses of eyes with and eyes without pigment plaque *de novo* formation. A total of 1216 eyes from 608 patients of 12 study centers were evaluated. 69 eyes from 69 patients (mean age 61.9 (range 53-71) years) were included and reviewed over a mean period of 41.6 months (range 24-60). 35 eyes (51%) showed a *de novo* development of pigmentary changes, and 34 eyes did not. Pigment plaques predominantly accumulated along vessels within the temporal parafovea (ETDRS subfield 5), usually sparing the fovea (Additional figure 1). Rarely, an extension of changes to the superior, inferior, or nasal parafovea (ETDRS subfields 2, 4, 3) was observed.

**Figure 1:**
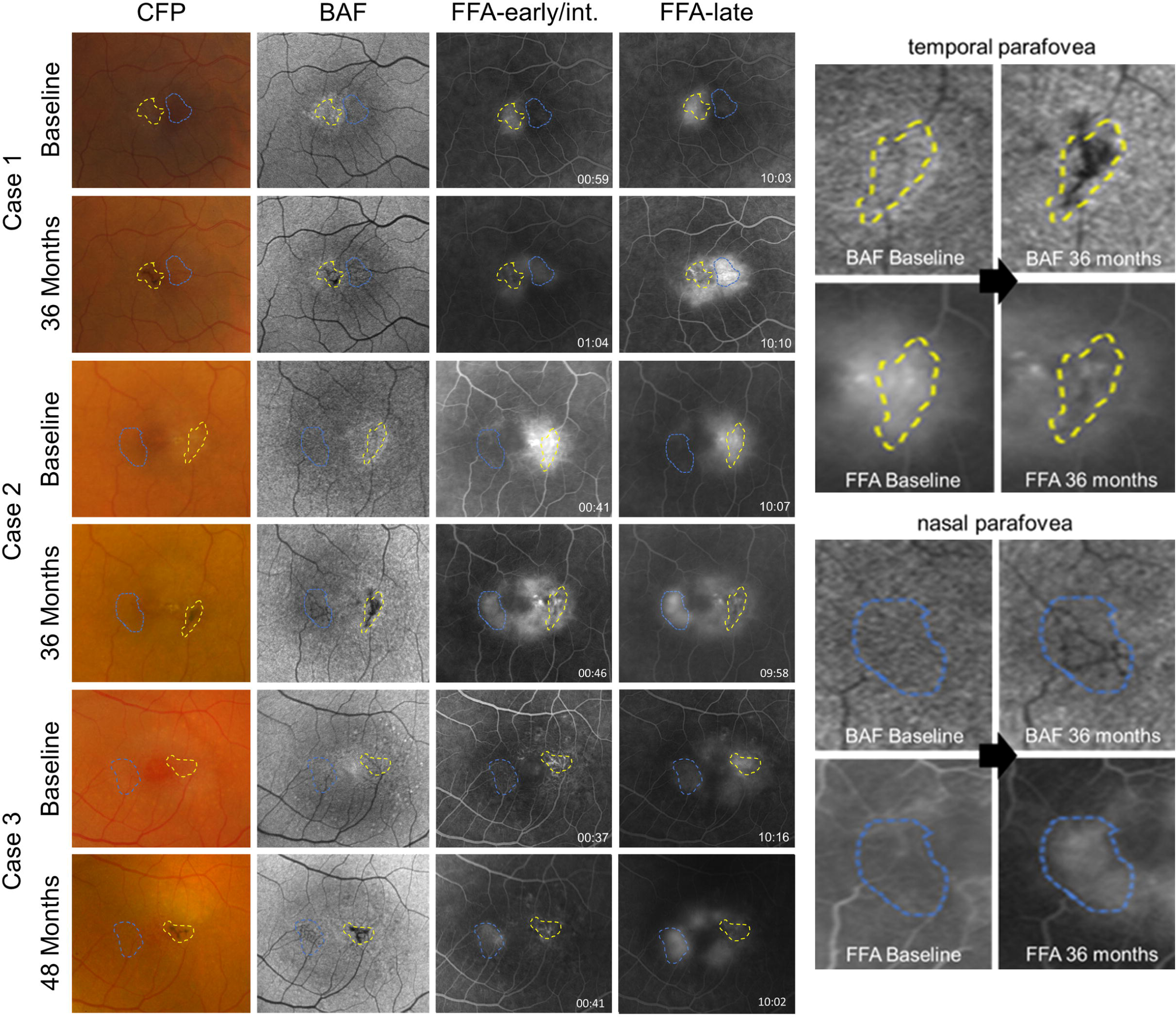
Perivascular accumulation of pigment plaques decreases vascular leakage in patients with MacTel. Longitudinal courses of three exemplary eyes showing a de novo formation of perivascular pigment plaques, imaged with color fundus photography (CFP), blue-light autofluorescence (BAF), and fundus fluorescein angiography (FFA; early to intermediate phase and late phase). A decrease in fluorescein leakage can be observed in vessels covered with pigment (yellow borderline), while vessels lacking pigment may show an increase in leakage and proliferation (blue borderline). The right column illustrates enlarged BAF and FFA images of case #2 within the temporal and nasal parafovea, respectively.

Longitudinal courses of eyes developing pigmented lesions differed from those without. A decrease in fluorescein leakage and stabilization of vessel proliferation was noted in all but one eye with pigment plaques. The observed effects were, however, focal and limited to vessels covered with pigment. In these eyes, vessels lacking pigment plaques showed stable, or rarely, increased leakage (observed in the nasal parafovea (ETDRS subfield 3) of 4/35 eyes; see figure 1). Notably, coverage of vessels with pigment was associated with a decrease in fluorescein leakage both in the early and late phase of fundus fluorescein angiography (FFA), suggesting a sealing rather than a mere shadowing effect associated with perivascular pigment accumulation. In eyes without pigmentary changes, an increase in vascular leakage (in 16/34 eyes [47%]) or stable leakage (in 18/34 eyes [53%]; table 1) was observed. Proliferation of vessels and increase in leakage were primarily observed within the temporal parafovea (ETDRS subfield 5). Exemplary longitudinal courses are illustrated in figures 1 and 2.

**Table 1:** Clinical findings in the study cohort of patients with macular telangiectasia type 2 (MacTel). *p-values were calculated based on an unpaired t-test with Welch’s correction. **p-values were calculated based on a Fisher’s exact test.

|  | **Pigment**<br>**plaques** | **No pigment**<br>**plaques** | **Statistical**<br>**significance** |
| --- | --- | --- | --- |
| **Eyes** | 35 [51%] | 34 [49%] | ns* |
| **Follow up period**<br>**(months; mean**<br>**[range])** | 41 [24-60] | 39 [24-60] | ns* |
| **Fluorescein leakage temporal parafovea (ETDRS subfield 5)** |  |  |  |
| **Increase** | 1 [3%] | 16 [47%] | p<0.0001** |
| **Decrease** | 34 [97%] | 0 | NA |
| **Stable** | 0 | 18 [53%] | NA |
| **Exudative**<br>**neovascularization** | 1 [3%] | 7 [21%] | p=0.0275** |
| **Visual acuity:**<br>**Loss of**<br>**letters/year (mean**<br>**[SD])** | 1.1 [2.3] | 3.1 [3.9] | p=0.0125* |

Exudative subretinal neovascularization is considered a severe, vision-threatening complication of MacTel, and is associated with severe vascular leakage. The de novo formation of exudative subretinal neovascular membranes was less frequently observed in eyes with compared to eyes without pigment plaques (in 1/35 [3%] eyes vs 7/34 [21%]; Fisher’s exact test, p=0.0275). Table 1 gives an overview of the clinical findings in this study cohort.

In summary, patients with MacTel showed (I) an accumulation of pigment plaques along abnormal retinal and subretinal vessels, (II) a decrease in vascular leakage that was associated with the development of pigment plaques, and (III) a decrease in exudative subretinal neovascularization associated with the presence of pigment plaques. Based on our findings in patients with MacTel, we hypothesized that (I) proliferating vessels may trigger the proliferation and perivascular accumulation of pigment. (II) Pigment plaques may be formed by RPE cells that undergo EMT, proliferate, migrate and accumulate along proliferating vessels. (III) Perivascular pigment plaques may decrease vascular leakage and stabilize vessel proliferation, thus having a beneficial effect on the diseased retina.

To test these hypotheses, and to further evaluate disease mechanisms leading to perivascular pigment accumulation, we studied related changes in the Vldlr-/- mouse model. Similar to eyes with MacTel, the Vldlr-/- mouse model shows a proliferation of retinal vessels, formation of retinal-choroidal anastomoses and subretinal neovascularization. With disease progression, RPE-cells proliferate and accumulate along subretinal neovessels, and subsequently migrate along retinal vessels into the neuroretina.[18, 19, 21]

### Proliferating retinal vessels trigger perivascular pigment accumulation

We first set out to investigate whether vascular proliferation triggers the proliferation and perivascular accumulation of pigment. In the Vldlr-/- mouse model, retinal vessels begin proliferating around P12, followed by the growth of retinal vessels to the outer retina, and the formation of subretinal neovascular complexes around P16-P21.[22] RPE-cells start proliferating around 4 weeks of age, accumulate along neovessels in the subretinal space, and subsequently migrate along retinal vessels into the neuroretina.[19, 21, 23] By inhibiting vascular proliferation using neutralizing antibodies against vascular endothelial growth factor (VEGF), we found a reduction in neovascular tuft formation. The ratio of pigmented to non-pigmented neovessels was, however, unchanged, and pigment plaques only developed along proliferating neovessels (see Figure 3A), suggesting that neovessels precede and are required for pigment plaque formation.

**Figure 2:**
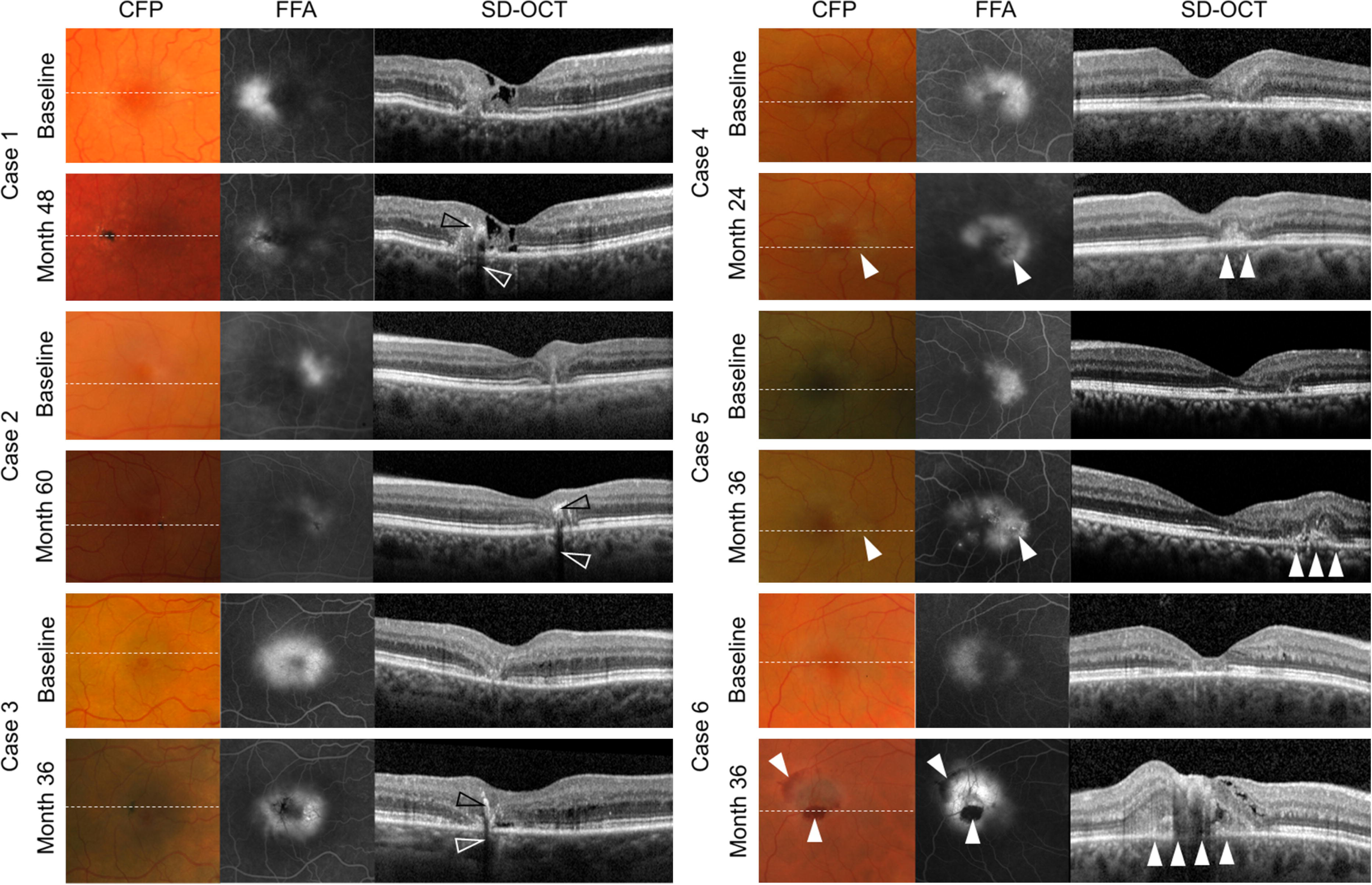
Perivascular pigment accumulation stabilizes vessel growth and decreases neovascular exudation in eyes with MacTel. Longitudinal courses of exemplary eyes with (cases 1-3) and without (cases 4-6) a *de novo* development of pigment plaques. Perivascular pigment accumulation is associated with a decrease in fluorescein leakage on fundus fluorescein angiography (FFA), and overall stable findings on spectral domain-optical coherence tomography (SD-OCT). Note the increase in intraretinal hyper-reflectivity (black arrowheads) with shadowing of underlying structures (white arrowheads) associated with pigment accumulation. Eyes without pigmentary changes show an increase in fluorescein leakage that may be associated with the *de novo* development of exudative neovascularization (white arrowheads). Hemorrhages may occur. CFP: color fundus photography.

**Figure 3:**
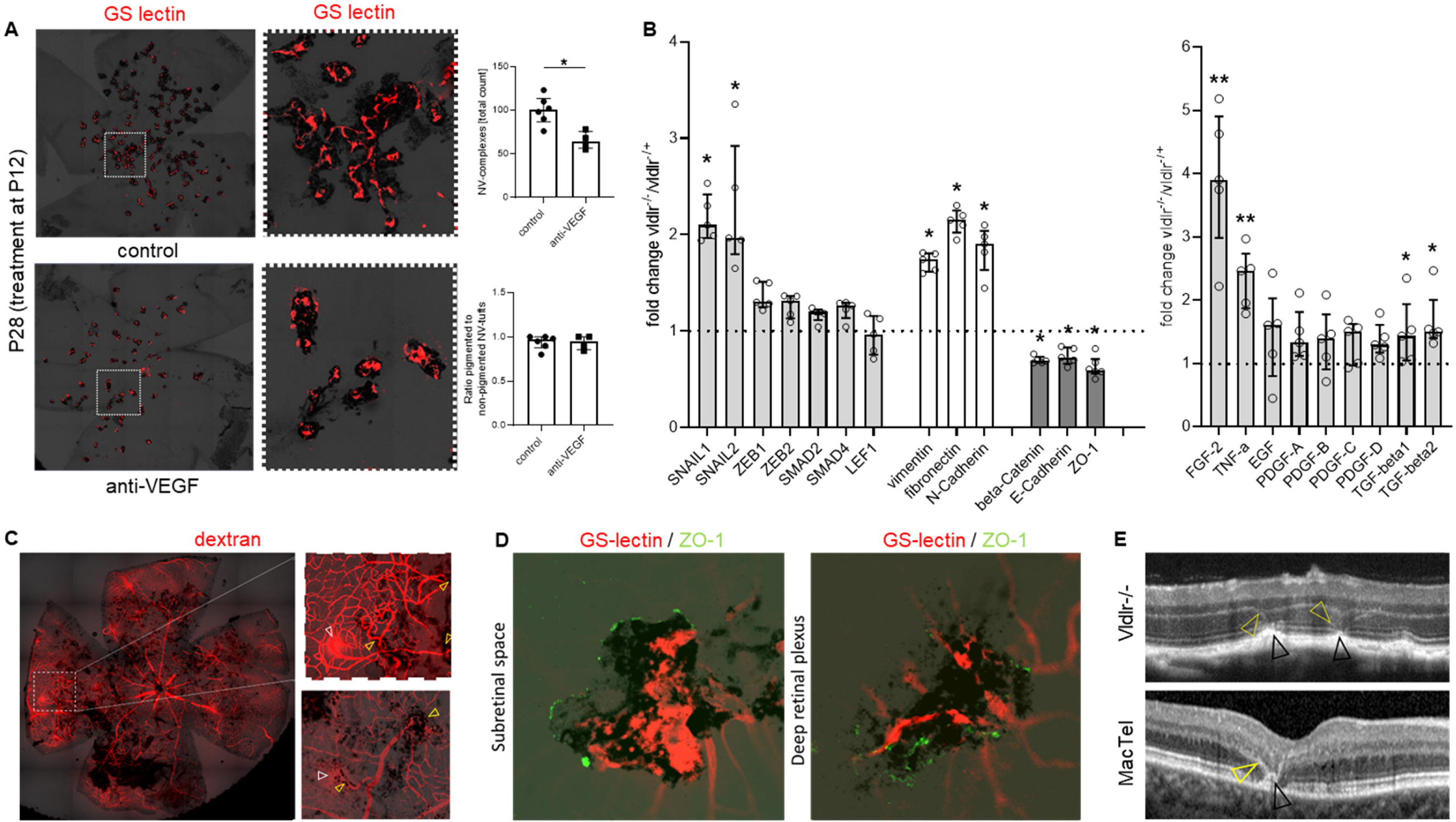
The Vldlr knockout mouse model mirrors vascular and pigmentary changes observed in MacTel. A: Proliferating vessels trigger perivascular pigment accumulation in Vldlr-/- mice. Neovascular (NV) tufts and perivascular pigment accumulation were analyzed using GS-lectin staining and bright field images in P28 Vldlr-/- mice treated with intravitreal injections of anti-VEGF (n=6) or control IgG (n=4) at P12. While anti-VEGF treatment significantly decreased the total number of NV-tufts (Mann-Whitney test, *p=0.019), the ratio of pigmented to non-pigmented NV-tufts was unchanged. Pigment plaques only developed along proliferating vessels. Error bars indicate the median and interquartile range. B: Regulation of different genes coding for key molecules and inducers of epithelial-mesenchymal transition (EMT) in the retina and RPE of Vldlr-/- mice at P42. Changes in genes between Vldlr–/– and Vldlr-/+ mice, as analyzed using a PCR array for EMT, are shown. P values were calculated based on a Mann-Whitney test of the replicate 2(-Delta Ct) values for each gene in the Vldlr-/- and Vldlr-/+ groups. *P < 0.05, **P < 0.01 (n=5 each). Median and IQR are shown for each gene. C: Dextran- angiography in a 12-months-old Vldlr-/- mouse depicts vessel and pigment proliferation and migration of pigmented cells along retinal vessels. Vessels covered with pigment show reduced dextran leakage (enlarged images, yellow arrowheads) compared to vessels not covered with pigment (white arrowheads). D: Pigment plaques cover proliferating vessels (stained with GS-lectin) and express ZO-1, indicating the formation of tight junctions along subretinal and retinal vessels in eyes of 10-months-old Vldlr-/- mice. E: Vldlr knockout (vldlr-/-) mice show hyper-reflective changes at the level of the outer retina (yellow arrowheads)/ retinal pigment epithelium (RPE; black arrowheads) on optical coherence tomography that resemble alterations observed in MacTel.

### Pigment plaques are formed by RPE cells in Vldlr-/- retinas and express similar markers as observed in eyes with MacTel

Previous findings in postmortem retinal samples of eyes with MacTel or retinitis pigmentosa indicated that intraretinal pigment plaques originated from the RPE.[3] Intraretinal lesions were found to express the epithelial cell marker cytokeratin-18 (CK- 18), that is specific to RPE-cells in the retina, but were negative for RPE65. Markers for mesenchymal cells (alpha-smooth muscle actin [a-SMA]) and macrophages/ microglia (IBA1) were also evaluated, but found to be absent.[3] To verify these findings in the Vldlr-/- mouse model, we evaluated similar markers as previously described.[3] Proliferating RPE cells within the subretinal space expressed CK-18 and RPE65. Some, but not all of these cells also showed immunoreactivity for a-SMA (see Additional figure 2), indicating EMT of the RPE. Intraretinal pigment plaques, on the other hand, expressed CK-18, but neither RPE65 nor IBA1 were detected (Additional figures 2 and 3). The expression of a-SMA was observed in single intraretinal pigmented lesions. The latter were, however, overall smaller and less dense compared to lesions lacking a-SMA expression, indicating a transitional, possibly less mature stage of these lesions (Additional figure 2).

### RPE-cells undergo EMT in Vldlr-/- retinas

Under physiologic conditions, the RPE is formed by a monolayer of polarized cells. Disintegration of the RPE monolayer and proliferation and migration of RPE-cells have been described in different degenerative retinal diseases, and have been attributed to RPE cells transitioning from an epithelial to a mesenchymal state.[7] To test whether RPE cells underwent EMT in the Vldlr-/- mouse model, we compared gene expression levels of known EMT-related genes in the RPE of Vldlr–/– mice and control Vldlr-/+ littermates at P42. At this timepoint, RPE cells have been shown to proliferate and accumulate along subretinal neovessels and start migrating along retinal vessels into the neuroretina.[18, 19, 21] Using qPCR arrays, we found an enrichment of genes coding for EMT pathways (SNAIL1/2) and different mesenchymal markers (vimentin, fibronectin, N-cadherin) in the RPE of Vldlr-/- mice. Genes coding for epithelial markers (beta-catenin, E-cadherin, zonula occludens-1 [ZO-1]), on the other hand, were decreased (Figure 3B), indicating that RPE cells underwent EMT in this model.

Next, we tested mRNA expression levels of known inducers of EMT in the retinas and RPE of Vldlr-/- mice. The highest differences between Vldlr–/– and heterozygous control littermates were found in fibroblast growth factor-2 (FGF2), which was increased 4-fold, and in tumor-necrosis factor-alpha (TNFa), which was increased 2.5-fold. FGF2 is a known driver of EMT that, among other factors, has been described to play a role in inducing EMT in RPE cells in proliferative vitreoretinopathy (PVR)[24]. FGF2 is also known to play a role in inducing subretinal fibrosis, and has been shown to have pro-angiogenic properties.[25, 26] TNFa is a proinflammatory cytokine and a known inducer of EMT in RPE cells.[7, 27] Elevated levels of TNFa have been detected in vitreous samples and epiretinal membranes of patients with PVR[28].[27, 29] *In vitro*, TNFa has been shown to induce RPE cells to upregulate EMT markers and mesenchymal key molecules.[30] Increased expression levels of TNF have previously been found in the retinas of Vldlr-/- mice, and in particular at the level of the deep retinal plexus.[22]

### Perivascular pigment decreases neovascular leakage and proliferation in Vldlr-/- retinas

Similar to eyes with MacTel, we found that in the Vldlr-/- mouse model vessels covered with pigment showed reduced dextran leakage compared to vessels not covered with pigment (Figure 3C). Perivascular pigment plaques expressed zonula occludens-1 (ZO-1), indicating the formation of tight junctions around proliferating vessels, thereby possibly reducing vascular leakage (Figure 3D). Furthermore, on OCT, Vldlr-/- mice showed hyper-reflective changes at the level of the outer retina/ RPE that resemble alterations observed in MacTel (Figure 3-E), which have been proposed to represent outer retinal neovascularization and proliferating RPE-cells.[17] Next, we set out to investigate whether inhibiting EMT of the RPE may impact neovascular leakage and proliferation in the Vldlr–/– model. Mice treated with neutralizing antibodies against FGF2 or TNFa from P21 to P42 showed a significant increase in vascular leakage and in the size of neovascular complexes at P42 in comparison to IgG-treated control animals (Figure 4A-C). Vascular leakage and perivascular pigment accumulation showed a negative correlation in both treated and control animals (Figure 4D).

**Figure 4:**
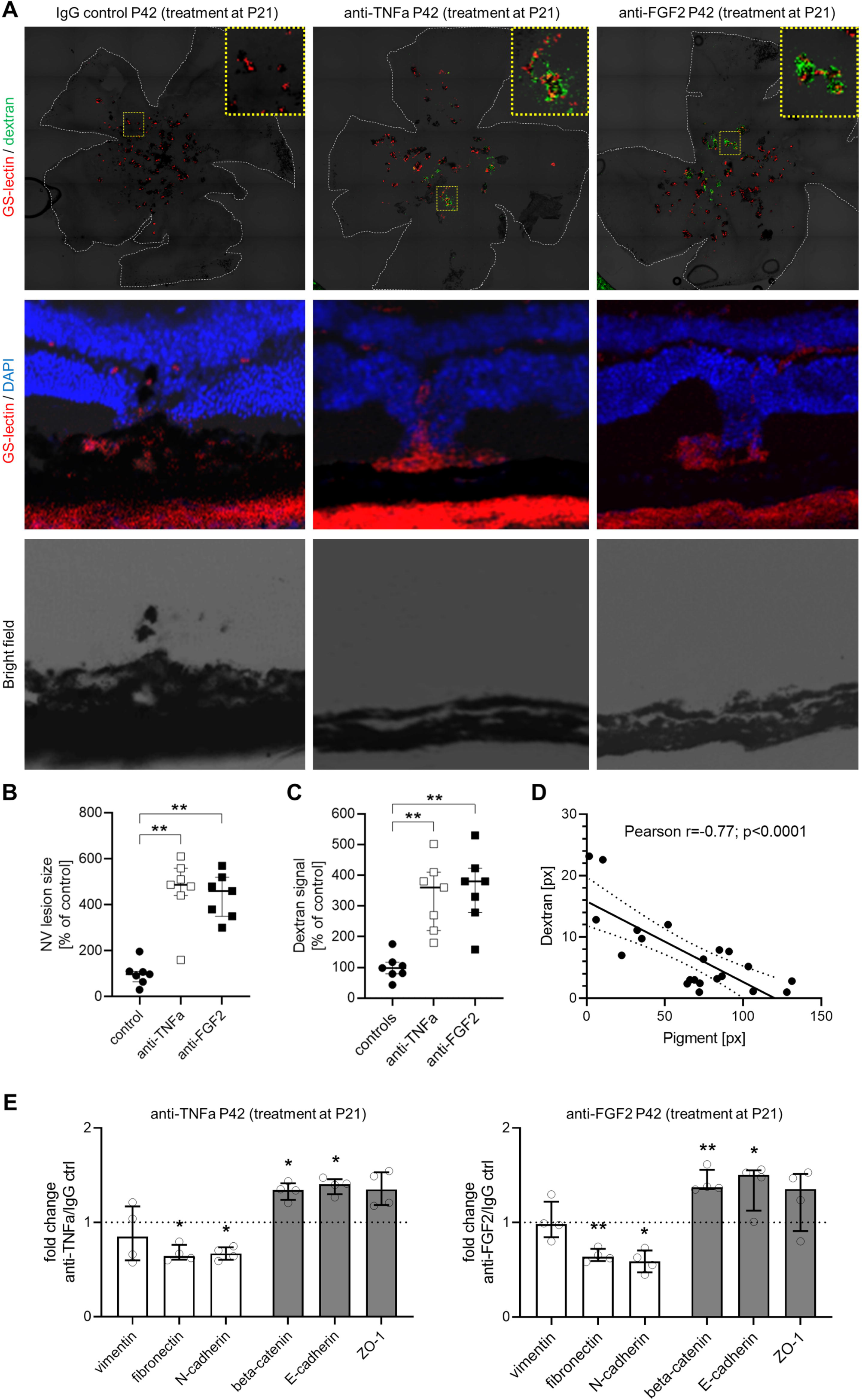
Inhibition of epithelial-mesenchymal-transition (EMT) leads to enhanced neovascular proliferation and leak in the Vldlr-/- model. A-C: Dextran angiography and GS lectin staining in flat mounted (A, upper panel) or cryo-sectioned (A, middle and lower panel) Vldlr-/- mice treated with intravitreal injections of neutralizing antibodies against TNFa (n=7) or FGF2 (n=7) at P21 showed a significant increase in dextran leakage (A, upper panel; C), and a significant increase in size of neovascular (NV) complexes (B) at P42 compared to IgG-treated controls (n=7). Yellow-dotted boxes in A (upper panel) show enlarged neovascular complexes in treated eyes. GS-lectin staining and bright field images of cryo-sectioned Vldlr-/- (A, middle and lower panel) illustrate decreased pigment proliferation and perivascular accumulation in eyes treated with anti-TNFa or anti-FGF2. D: Dextran-leakage (number of pixels positive for dextran) was negatively correlated (Pearson r=-0.78; p<0.0001) with pigment accumulation (number of pixels positive for pigment) in flat-mounted Vldlr-/- eyes. E and F: Changes in genes coding for key molecules of EMT in Vldlr–/– mice at P42 treated with either anti-TNFa (E) or anti-FGF2 (F) compared to IgG-treated Vldlr-/- mice (treatment at P21), as analyzed using a PCR array for EMT, are shown. While mesenchymal key molecules (white bars) were downregulated, epithelial key molecules were enriched (grey bars). P values were calculated based on a Mann-Whitney test of the replicate 2(-Delta Ct) values for each gene. *P < 0.05, **P < 0.01 (n = 4 each).

To test whether the observed morphological changes were associated with the inhibition of EMT, we compared gene expression levels of EMT-related genes in the RPE of Vldlr–/– mice treated with FGF2, TNFa or control IgG. While no changes were observed for EMT pathways, genes coding for epithelial markers were enriched, and mesenchymal markers were decreased in animals treated with FGF2 or TNFa, indicating the inhibition of EMT (Figure 4E).

In summary, we suggest that the perivascular accumulation of RPE-cells may stabilize neovascular proliferation and leakage, thereby exerting a beneficial, protective effect on the diseased retina. Figure 5 summarizes the herein proposed mechanisms in a schematic illustration.

**Figure 5:**
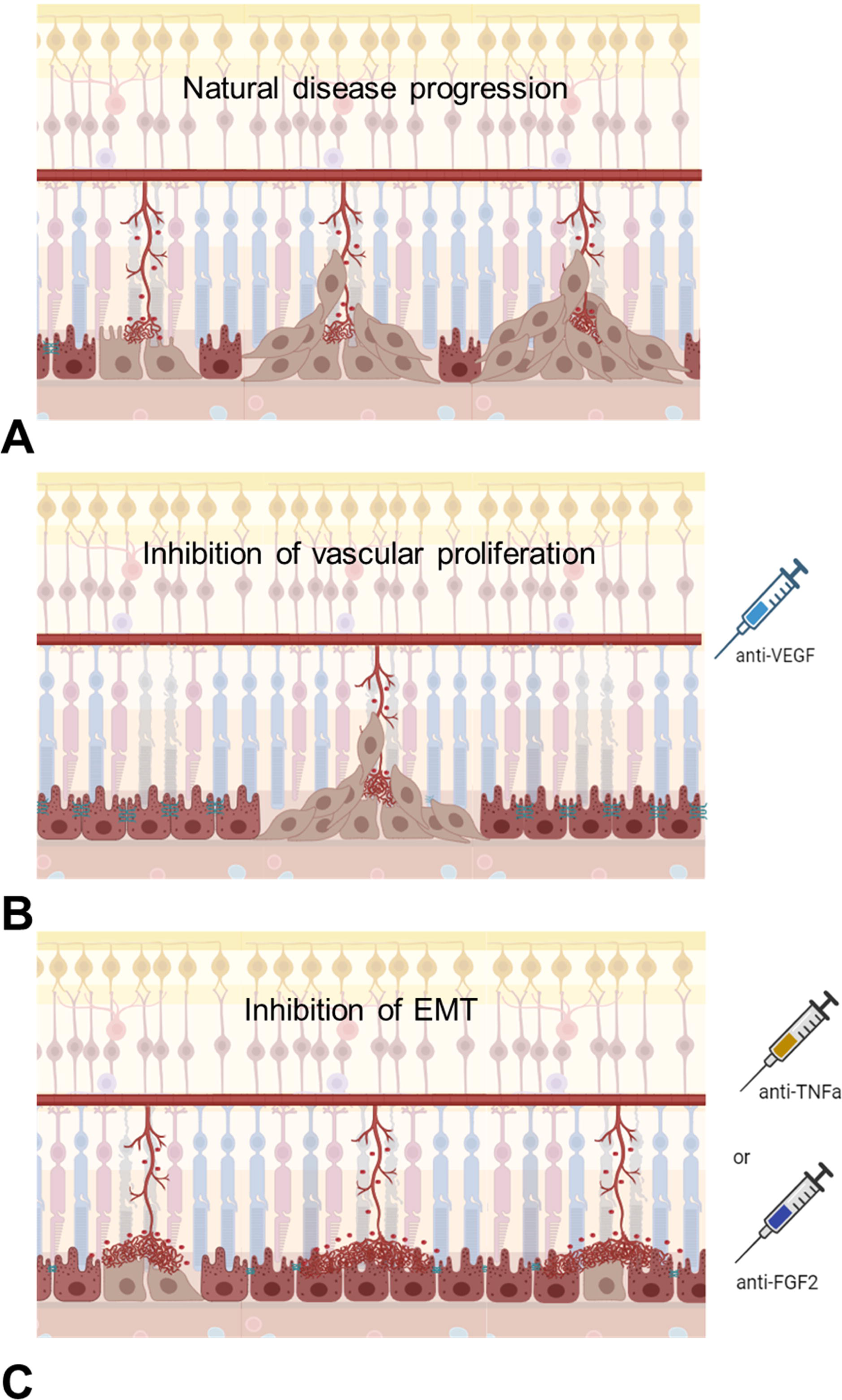
Schematic illustration of the proposed interplay between RPE-cells and retinal vessels in the Vldlr-/- model. A: Proliferating vessels of the deep retinal plexus grow to the outer retina, form neovascular tufts and get in contact with the RPE. RPE-cells transition from an epithelial to a mesenchymal state, proliferate and migrate along retinal vessels into the neuroretina, forming perivascular plaques. B: Eyes treated with intravitreal antibodies against VEGF (anti-VEGF) show a reduction in vessel proliferation and decreased numbers of neovascular tufts, while EMT of the RPE is not impacted. C: In eyes treated with intravitreal antibodies against TNFa or FGF2, epithelial-mesenchymal transition (EMT) of the RPE is inhibited. RPE cells show reduced proliferation, perivascular accumulation and migration. Neovascular tufts show more leakage and increased lateral growth. Created with BioRender.com

## Discussion

Pigment plaques can be found in different retinal diseases involving vascular pathology and neurodegeneration. The origin of these changes has been attributed to RPE-cells, accumulating along subretinal and retinal vessels, and migrating along vascular structures into the neuroretina.[3, 20] The role these changes may play during disease progression is, however, not yet understood. In this study, we sought to evaluate the impact of perivascular pigment plaques on the progression of vascular changes in eyes with MacTel. We observed an association of perivascular pigment with a focal decrease in vessel permeability, and stabilization of vessel proliferation. The development of exudative subretinal neovascular complexes was observed less frequently in eyes with compared to eyes without pigment plaque formation. In the literature, differing numbers describing potential associations of pigment plaques with subretinal neovascularization in MacTel have been reported. While Leung et al. found that in about 25% of neovascular eyes, subretinal neovascularization coincided with or preceded pigmentary changes[5], Engelbrecht et al observed that pigmented lesions preceded the development of neovascular membranes in all cases (in 11/11 eyes).[14] Meleth et al reported the *de novo*-development of neovascularization in equal numbers of eyes with and without preexistent pigmentations, respectively.[31] Differences between our data and previous reports may be explained by differing definitions of “neovascularization” and “exudation”, based on distinct imaging modalities, and differing in- and exclusion criteria, resulting in study populations with varying disease stages, baseline characteristics and observational periods. More recent studies evaluating longitudinal OCT-A data found that the formation of outer retinal neovascularization preceded perivascular pigment accumulation in all cases.[32] Based on their findings on multimodal retinal imaging, Mueller et al suggested that outer retinal neovascularization induced proliferation of the RPE, once retinal vessels get in contact with the RPE[17], a finding that is similar to previous observations in Vldlr-/- mice and other models.[18, 19, 21, 33] Our results are in line with these later findings of neovascularization preceding pigmentation and indicate that perivascular pigment accumulation is a focal process, possibly triggered by vascular leakage and proliferation, and may represent a “repair mechanism” of the disrupted RPE. Furthermore, our results may indicate that pigment plaques could be used as a biomarker to predict neovascular exudation and vascular stabilization, respectively. Prospective, longitudinal studies are needed to verify our findings.

The reason why only a subset of MacTel patients develop pigment plaques currently remains obscure and warrants further studies. Limited observational periods and patient numbers might have confounded our findings, representing potential limitations of this study.

To study underlying disease mechanisms, we evaluated related pigmentary and vascular changes in the Vldlr-/- mouse model. We showed that perivascular pigment plaques expressed similar markers as previously described in postmortem retinal specimens of eyes with MacTel and other diseases, verifying that these changes are formed by RPE-cells.[3, 20] We confirmed that pigment accumulation was associated with a decrease in vessel leakage in the Vldlr-/- model. Our findings indicate that perivascular pigment plaques may form tight junctions around proliferating vessels, as indicated by ZO-1 localization, thereby reducing vascular leakage and stabilizing neovascular growth. A similar effect of pigment plaques on vessel permeability has been previously reported by Jaissle et al, where migrating RPE-cells were observed to form tight junctions with vascular endothelial cells, thus sealing retinal vessels in a mouse model for retinitis pigmentosa.[33]

While dense pigment plaques within the neuroretina and subretinal space of Vldlr-/- mice showed a focal expression of ZO-1, genes coding for the same epithelial cell marker were downregulated in the RPE of Vldlr-/- mice, indicating a broader loss of cell-cell contacts and epithelial cell state. This suggests a loss of adhesion in the general RPE monolayer, perhaps indicative of cells undergoing transition to a more migratory phenotype. Taken together, these findings might indicate that RPE-cells undergo EMT, proliferate and accumulate along subretinal and retinal vessels. Once migrating RPE-cells have found their new position, they may develop cell-cell contacts with neighboring RPE-cells, forming dense plaques around the vessels, and possibly transition back to an epithelial state (“mesenchymal-epithelial transition”, MET). Similar mechanisms have been previously described to occur during cancer metastasis and organ morphogenesis.[34, 35] The hypothesis that RPE cells undergo EMT, followed by MET, in the Vldlr-/- model is further supported by the differences we found in the expression of epithelial and mesenchymal markers in Vldlr-/- mice compared to previous reports in MacTel. In MacTel, intraretinal pigment plaques were found to only express the epithelial marker CK18, while lacking any mesenchymal markers.[3] In the Vldlr-/- model, on the other hand, we observed that single intraretinal pigmented lesions also expressed mesenchymal markers, possibly indicating the presence of different stages of lesions. This was particularly the case for lesions that were less dense, and thus possibly less mature, indicating that some RPE-cells were still in a mesenchymal state, before undergoing MET and forming dense, mature plaques around the vessels. In MacTel, on the other hand, intraretinal pigment plaques usually develop within months and may form and progress over the course of several years, forming dense, organized clusters of pigment around vessels.[3, 5] Thus, it is conceivable that lesions, once clinically detectable, are overall more mature, and RPE-cells within dense plaques no longer possess mesenchymal properties, possibly having transitioned back to an epithelial state. Future studies are warranted to further explore the proposed mechanisms.

EMT of RPE-cells has been described in different ocular conditions[7] and various inducers of EMT have previously been discussed.[7, 25, 29, 30] In the RPE of Vldlr-/- mice, we found several known drivers of EMT to be enriched, with FGF2 and TNFa being the most significant ones in our model. While these factors could be related to general neovascularization events in the Vldlr-/- retina, it is important to note that their inhibition actually reduced neovascularization and leakage, thus indicating a role in EMT rather than general neovascularization. Other known drivers of EMT and/or fibrosis have previously been identified in the Vldlr-/- model.[36, 37] Notably, EMT and TNFa/ NFκB pathways were previously found to be among the most significantly enriched gene sets in RPE cells differentiated from induced pluripotent stem cells from MacTel donors.[38] This underlines the clinical relevance and applicability of our findings in the Vldlr-/- model.

Inhibiting EMT in the Vldlr-/- model resulted in increased neovascular growth and exudation, confirming the beneficial effect of perivascular pigment accumulation we had clinically observed in patients with MacTel. Pigment plaques, on the other hand, have been shown to be associated with a focal loss in retinal sensitivity (“absolute scotomas” on fundus-controlled perimetry)[8, 39] and the pigment area with a decrease in visual acuity in patients with MacTel[5], thus representing a mixed blessing.

Intravitreal VEGF inhibitors have been successfully used to treat numerous retinal diseases showing vascular involvement. In MacTel, VEGF inhibitors have been effectively applied in eyes showing subretinal neovascularization[40], while no functional improvement has been reached in non-neovascular eyes.[41] The latter showed, however, a (temporary) decrease in fluorescein leakage during treatment, followed by an increase in leakage once treatment had been discontinued.[41] Notably, treated eyes developed excessive pigmentation and fibrosis later on.[41, 42] In the Vldlr-/- model, we found that perivascular pigment only developed when vascular proliferation was present and pigment plaques only developed along proliferating vessels. Taken together, this might further indicate that interfering with the herein proposed natural “repair mechanism” of the diseased retina/RPE, by suppressing either EMT or vascular leakage/ intraretinal proliferation, may have detrimental effects and thus should be avoided in MacTel and related diseases.

## Conclusions

In summary, we showed that perivascular pigment accumulation is associated with a decrease in vascular leakage and stabilization of neovascular growth in proliferative retinal diseases, such as MacTel, and respective animal models. We revealed underlying mechanisms leading to perivascular pigment accumulation, and discussed beneficial effects, changing our current understanding of these changes. Knowledge about pathophysiological mechanisms is crucial for understanding the disease course and developing therapeutic interventions as well as choosing appropriate timepoints for treatment. We conclude that interfering with this “natural repair mechanism” of the diseased RPE may have detrimental effects on the course of the disease and should thus be avoided.

## Materials and Methods

### Participants

In a retrospective, longitudinal approach, imaging data of affected participants from twelve participating sites of the multi-center Natural History Study of Macular Telangiectasia (MacTel Study) were evaluated. Protocol details of this study have been published previously.[43] The diagnosis of MacTel type 2 was based on characteristic morphologic findings on fundoscopy, OCT, fundus autofluorescence and fluorescein angiography[8], and was confirmed by the Moorfields Eye Hospital Reading Centre, London, UK. Patients underwent annual study visits, and eyes were reviewed over a minimum observational period of 24 months. The studies were approved by the local ethics committees at each participating study site and were in adherence with the Declaration of Helsinki. All participants provided informed consent. Data were collected at a minimum of two time points (at baseline and last available follow up visits), and sites were selected based on the availability of longitudinal imaging data, including color fundus photography (CFP), Spectral Domain-OCT (SD-OCT; volume scans of 15° x 10° (high resolution mode, 97 scans) or 25° x 30° (high speed mode, 61 scans); Spectralis, Heidelberg Engineering, Heidelberg, Germany), fundus autofluorescence (FA) and fundus fluorescein angiography (FFA; 30°, centered on the fovea, images taken at 30 seconds, 1, 5 and 10 minutes after fluorescein injection). In a small subset of participants, OCT-angiography (OCT-A) data were additionally available.

Inclusion criteria for this analysis were a confirmed diagnosis of MacTel, a full data set including the examinations listed above and sufficient image quality. As pigmentary changes have been shown to only develop in eyes showing disruptions of outer retinal layers[44], and to ensure equal baseline conditions among both groups, only eyes with intermediate disease stages were included. The latter was defined as visible disruption of the ellipsoid zone (EZ)/ photoreceptor layer on OCT, and disease stages 2-3 according to Gass and Blodi.[13] Eyes showing pigment plaques and/ or subretinal/ sub-RPE neovascular membranes (stages 4 and 5 according to Gass and Blodi) at baseline were excluded. Further exclusion criteria were the presence of potentially confounding retinal diseases including central serous chorioretinopathy, age-related macular degeneration or diabetic retinopathy, and previous therapies including VEGF-inhibitors, vitreo-retinal surgery, photodynamic therapy or central laser treatment.

Eyes were retrospectively divided into two groups, one group showing a *de novo* formation of pigment plaques at last follow up, and one group without any pigmentary changes, and outcome parameters were compared between both groups.

Outcome parameters included changes in leakage on FFA (decreased, increased, or stable leakage within subfields 1-5 of the ETDRS grid) and a *de novo* development of exudative subretinal neovascular membranes as observed on CFP, OCT, FFA, and OCT-A (if available).

### Image grading and definitions

### Pigment plaques

CFP and FA images were analyzed for the presence and position of pigment plaques at baseline and last follow up.

### Leakage on FFA

On FFA, vessels were graded for an (1) increase, (2) decrease, or (3) no visible changes in fluorescein leakage at last follow up compared to baseline. Images were evaluated and compared at different time points (at 30 seconds, 1 minute, and 10 minutes after fluorescein injection) and within subfields 1-5 of the ETDRS grid.

### Neovascularization and exudation

Previously described criteria were applied to identify neovascular membranes on OCT, FFA, CFP, and OCT-A images (if available).[16, 44] Neovascular exudation was defined as focal retinal thickening, sub- or intraretinal fluid, and/or hemorrhages as observed on OCT and/ or CFP in neovascular eyes.

### Animals

All animal experimental procedures were approved by The Scripps Research Institute Animal Care and Use Committee. Experiments were performed in accordance with the NIH Guide for the Care and Use of Laboratory Animals (National Academies Press, 2011). Vldlr–/– mice (15) and control littermates (Vldlr-/+ mice) (The Jackson Laboratory) of up to 12 months of age were used for all animal experiments.

### Intravitreal injections

All intravitreal injections were performed using a Hamilton syringe and a 34-gauge needle (Hamilton), injecting 1μl of solutions containing neutralizing antibodies against TNFa (250ng; MAB4101; R&D systems), FGF2 (200ng; clone bFM-1; 05-117, Millipore Sigma) or VEGF164 (200ng; AF-493-NA; R&D systems). For each of these antibodies, IgG isotype control antibodies were used as negative controls (MAB005R, R&D systems; 12-371, Millipore Sigma; AB-108-C, R&D systems).

Intravitreal injections with antibodies against FGF2 or TNFa were performed at P21, after subretinal neovascular tufts have formed, but before pigmented cells have started proliferating or migrating[18, 45]. Retinas and RPE/choroid/sclera complexes were analyzed for gene expression or immunofluorescence at P42. At this timepoint, RPE-cells have been shown to proliferate and accumulate along subretinal neovessels and have started migrating into the neuroretina, where they accumulate along retinal vessels.[18, 19, 21, 45]

Intravitreal injections with antibodies against VEGF were performed at P12, when retinal vessels have started proliferating and anti-VEGF treatment has been shown to be most effective in this model.[21, 36] Retinas were evaluated at P28 using immunofluorescence imaging. At this timepoint, the normal RPE monolayer is disrupted, and clumps of RPE cells accumulate along neovascular tufts in the subretinal space of Vldlr-/- mice.[19, 21]

### Dextran angiography

For analyzing vascular leakage, Fluorescein isothiocyanate– dextran (FITC; 40,000 MW; Sigma-Aldrich) was perfused through the left ventricle of deeply anesthetized Vldlr-/- animals using 150μl of a 50mg/ml solution in PBS.

### Immunofluorescence

Retinas and RPE/choroid complexes were dissected and prepared for whole mounts or sectioning. For preparation of retinal cross-sections, isolated eyes were fixed in 4% paraformaldehyde (PFA) for 4 hours, placed in 15% sucrose overnight at 4°C, followed by 30% sucrose for 2 hours, and embedded in Tissue-Tek OCT compound (Sakura Finetek) for subsequent cryosectioning. For preparation of whole mounts, eyes were fixed in 4% PFA for one hour, and retina/RPE/choroid complexes were dissected and laid flat. Whole-mount retina/RPE/choroid complexes or cryosections were incubated in blocking buffer (10% fetal bovine serum and 0.1% Triton X-100 in phosphate-buffered saline [PBS]) at 4°C for 2 hours, followed by an incubation with primary antibodies in blocking buffer at 4°C overnight. Specimens were then washed with PBS and incubated with Alexa Fluor– conjugated secondary antibodies (Thermo Fisher) for 2 hours. Nuclei were stained using DAPI (Vector Laboratories). Specimen were mounted in Vectashield Plus Antifade Mounting Medium (Vectorlabs). Primary antibodies against ZO-1 (40-2200, Thermo Fisher), RPE-65 (PA5-110315, Thermo Fisher), alpha-smooth muscle actin (ab124964, Abcam), IBA1 (MA5-27726, Thermo Fisher) and Cytokeratin 18 (10830-1-AP, Proteintech) were used. Endothelial cells were labeled using Fluorescent-conjugated isolectin Griffonia Simplicifolia IB-4 (GS-Lectin) (I21412, Thermo Fisher). Images were captured with a confocal laser scanning microscope (LSM 710, Zeiss) and processed with the ZEN 2010 software (Zeiss). The NIH ImageJ software was used for quantifying dextran-leakage (number of pixels positive for dextran), pigment accumulation (number of pixels positive for pigment) and neovascular tufts (total numbers of visible tufts per eye).

### Gene expression analyses

For quantitative polymerase chain reaction (qPCR), retinas and RPE/choroid complexes were isolated in 500μl Trizol. RNA isolation was performed using a RNeasy Plus Micro Kit (Qiagen) following the manufacturer’s instructions. 400ng RNA was used for real time-qPCR using the High-capacity cDNA Reverse transcriptase kit (Thermo Fisher). SYBR Green–based (Thermo Fisher) real-time quantitative PCR was performed on a QuantStudio 5 (Thermo Fisher) to analyze mRNA expression levels of various gene products. Expression levels were normalized to Cyclophilin A. Sequences of primers used are listed in supplementary table 1. Data were analyzed using the QuantStudio Design and Analysis Software 2.6.0 (Thermo Fisher).

### Statistical analysis

Statistical analyses were performed using GraphPad Prism, version 10.1.0 (GraphPad Software, San Diego, CA, USA). Continuous variables were described by using the mean ± standard deviation (SD), median and interquartile range [IQR] and/ or ranges. Categorical variables were described in terms of frequency. Unpaired t-test with Welch’s correction was applied for comparing parameters between two groups in normally distributed samples. Mann–Whitney test was used to compare parameters between two groups in non-normally distributed samples. Kruskal-Wallis test with Dunn’s correction for multiple comparisons was used to compare data between multiple groups. Fisher’s exact test was applied to analyze contingency tables with small sample sizes. Pearson correlation coefficient was calculated to determine linear correlation between two parameters. Statistical tests applied for each experiment are detailed in the figure and table legends. A p-value <0.05 was accepted as statistically significant.

## Data Availability

All data produced in the present work are contained in the manuscript.

## Supplement

Supplementary Figure 1: Pigment accumulates along proliferating retinal vessels and retinal-choroidal anastomoses in eyes with MacTel.

Multimodal retinal images of four exemplary eyes with MacTel, including optical coherence tomography-angiography (enface scans of the following layers are shown: whole retina, superficial retinal plexus, deep retinal plexus and whole eye scans; vessels forming retinal-retinal and retinal-choroidal anastomoses are marked in red), B-scan Spectral domain (SD-) OCT, color fundus photographs (CFP), blue-light autofluorescence (BAF), and fundus fluorescein angiograms (FFA, late phase). Pigment plaques are indicated with arrow heads. On OCT, pigment plaques depict as hyper-reflective lesions. Horizontal lines on CFP images indicate the position of each B-scan.

Supplementary Figure 2: Dense intraretinal pigment plaques express the retinal pigment epithelium marker Cytokeratin-18 (CK-18).

Immunofluorescence imaging of retinal sections of 11-months-old Vldlr-/- mice with GS-lectin (red) and CK-18, alpha-smooth muscle actin (a-SMA), IBA-1 or RPE65 (green). Nuclei are shown in blue (DAPI). Intraretinal pigment co-locates with CK-18, but not with a-SMA, IBA1 or RPE65.

Supplementary Figure 3: RPE-cells form clusters around neovessels within the subretinal space, and express epithelial and mesenchymal markers.

Dense intraretinal pigment plaques (along vessels of the deep plexus) express the RPE marker Cytokeratin-18 (CK-18), but not RPE65. The expression of a-SMA was observed in single pigmented intraretinal lesions (black arrowhead). The latter appeared, however, overall less dense compared to those lesions lacking a-SMA expression (white arrowhead). Immunofluorescence imaging of flat mounted retinas of 12-months-old Vldlr-/- mice with GS-lectin (red) and CK-18, alpha-smooth muscle actin (a-SMA), IBA-1 or RPE65 (green). Nuclei are shown in blue (DAPI).

