## Supplementary figures and images for "Retinal pigment epithelial cells reduce vascular leak and proliferation in retinal neovessels"

### Supplemental Figure 1

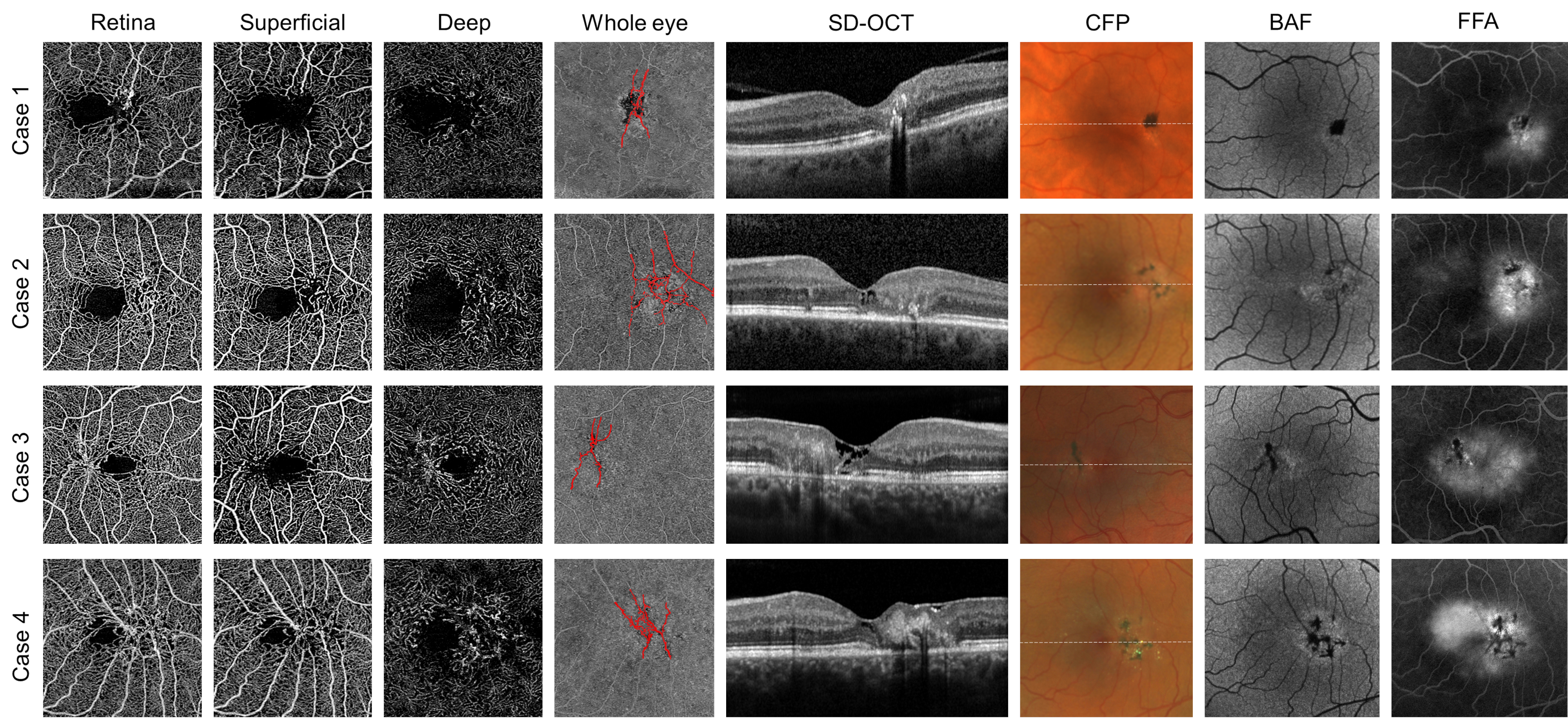

### Supplemental Figure 2

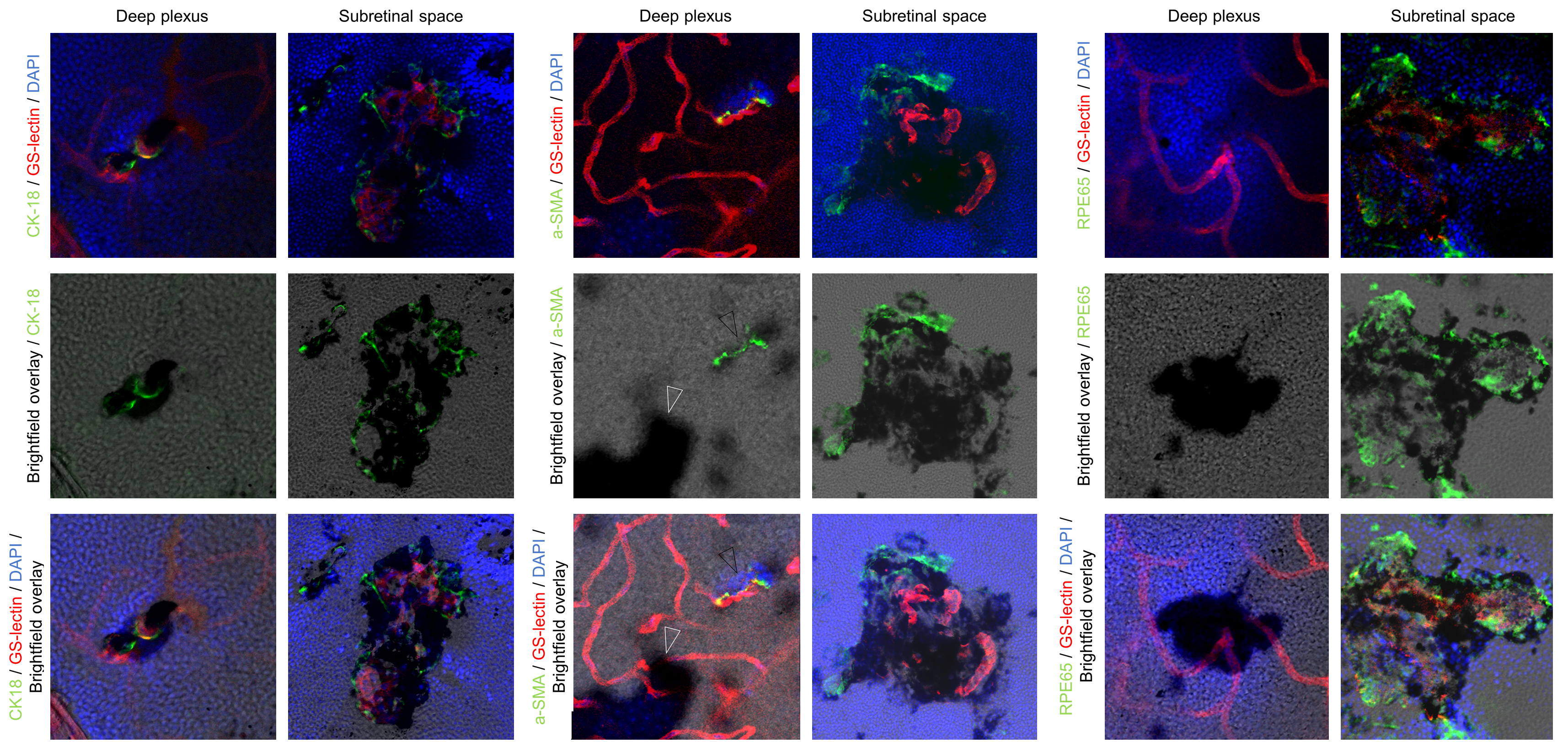

### Supplemental Figure 3

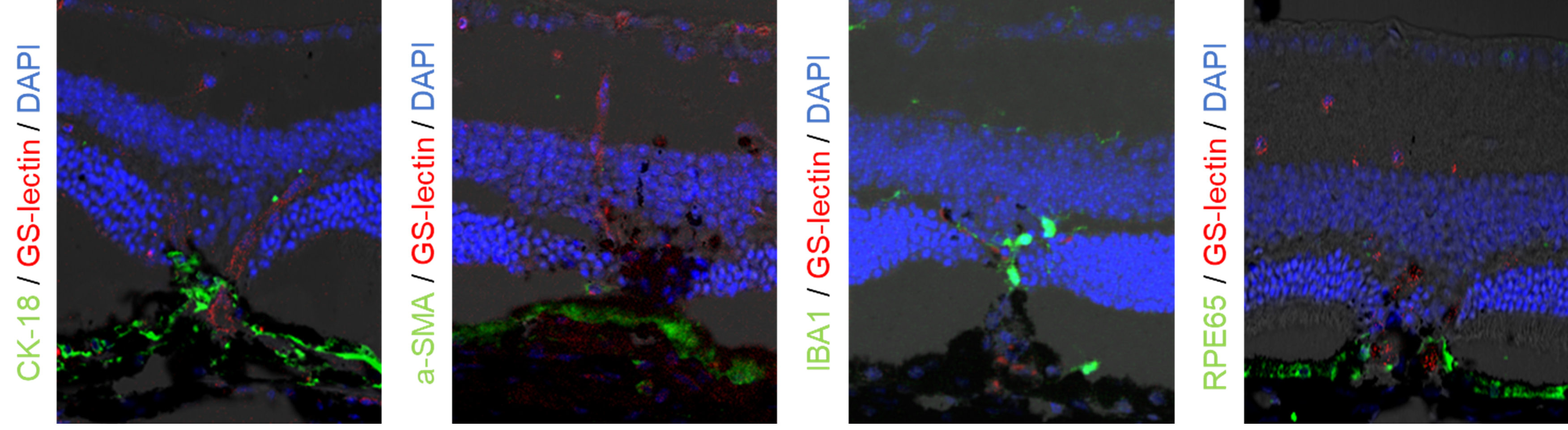
