## Supplemental Table 1 for "Retinal pigment epithelial cells reduce vascular leak and proliferation in retinal neovessels"

Additional table 1:

Sequence of primers used for RT-PCR.

| **Gene** | **Forward primer (5’-3’)** | **Reverse primer (5’-3’)** |
| --- | --- | --- |
| TNF-alpha | CCCTCACACTCAGATCATCTTCT | GCTACGACGTGGGCTACAG |
| FGF-2 | CAGTTCGTTTCAGTGCCACA | GGCTGCTGGCTTCTAAGTGT |
| EGF | ATTTCGTTGTTAGCACCATCCC | GGCACAACCAGGCAAAGGAT |
| TGF-beta-2 | TCCCCTCCGAAAATGCCATC | TGCTATCGATGTAGCGCTGG |
| N-cadherin | TCTCCAAGTGGCCAGGAAAC | CAAAGCTTCCGGGCGTAGA |
| ZO-1 | GACCTCTGCAGCAATAAAGCA | AGAAATCGTGCTGATGTGCCA |
| E-cadherin | AACCCAAGCACGTATCAGGG | GAGTGTTGGGGGCATCATCA |
| vimentin | GGCTGCGAGAGAAATTGCAG | TTCAAGGTCAAGACGTGCCA |
| SMAD2 | TTGCTGTTGTTGTTGTTTAAGGA | AGCAATACTGCCTCTTGTTGC |
| SMAD4 | AGTAATCGCGCATCAACGGA | GAATACTGGCCGGCTGACTT |
| ZEB1 | CTGAGCACAGACTACCGCAA | GGTCTGCTGGCAGTTCATCA |
| ZEB2 | AAGCGTTTGCGGAGACTTCA | AACACGCGCCACCTATCTTT |
| SNAIL1 | AGTTGACTACCGACCTTGCG | TGCAGCTCGCTATAGTTGGG |
| SNAIL2 | AGAAGCCCAACTACAGCGAA | ATAGGGCTGTATGCTCCCGA |
| beta-catenin | CGCCGCTTATAAATCGCTCC | TTCACAGGACACGAGCTGAC |
| LEF1 | CAGCGCGAGACAATTATGGC | TAGGCAGCTGTCATTCTGGG |
| fibronectin | CTGGATCCCCTCCCAGAGAA | TTGGGGTGTGGAAGGGTAAC |
| TGF-beta1 | ACTGGAGTTGTACGGCAGTG | GGGGCTGATCCCGTTGATTT |
| PDGF-A | TTCGTCGATAACACGCACGA | TTCCCAGAGTCCCCTCATGT |
| PDGF-B | CAACGAGAAAGCCGGAGCAG | GTCTATCTACCCACTCGCTCG |
| PDGF-C | CCAGTCAGCCAAATGCTCCT | TGGGTATAGTTCCTCCCGTTCT |
| PDGF-D | TGAGAGCAATCACCTCACAGAC | CAGAAGCAGGTTCCTTGGGT |
